# Tracking premenstrual exacerbation (PME) of depression in a prospective clinical cohort: the TIDE study protocol

**DOI:** 10.64898/2026.05.01.26352210

**Authors:** Chamee A. Giezenaar, Isis de Valk, Margot W. L. Morssinkhof

## Abstract

**Introduction:** There is a growing body of research showing that the menstrual cycle can affect mood, although research in those with an existing depressive disorder is still scarce. Studies estimate that 60% of women with depression experience premenstrual exacerbation (PME) of their depressive symptoms.

**Aims:** With the TIDE study, we aim to 1) examine the feasibility of daily symptom tracking for two consecutive menstrual cycles to track PME, 2) estimate the prevalence of PME in depression in a clinical cohort and 3) examine whether PME is associated with other hormone-related mood symptoms (i.e., hormonal contraceptive side effects, peripartum depression).

**Methods:** We aim to recruit 60 female participants aged 18 to 45, who are in treatment for a depressive episode and who have a regular natural menstrual cycle. Participants will participate in questionnaires at baseline, inquiring about demographic characteristics and previous experiences with hormonal contraceptives and pregnancy. Participants will complete retrospective menstruation questionnaires on days 1 and 10 of each menstrual cycle, as well as daily diaries for two consecutive menstrual cycles, inquiring about menstruation and mood symptoms. After completion of the diaries, participants will receive a symptom report, as well as a study evaluation questionnaire.

**Results and conclusion:** We expect that providing the menstrual cycle overview of symptom severity will lead to increased symptom course insights in participants with PME, and that participants with PME will find it clinically relevant and significant to gain insight into symptom trajectories across the menstrual cycle.

**Graphical abstract:** 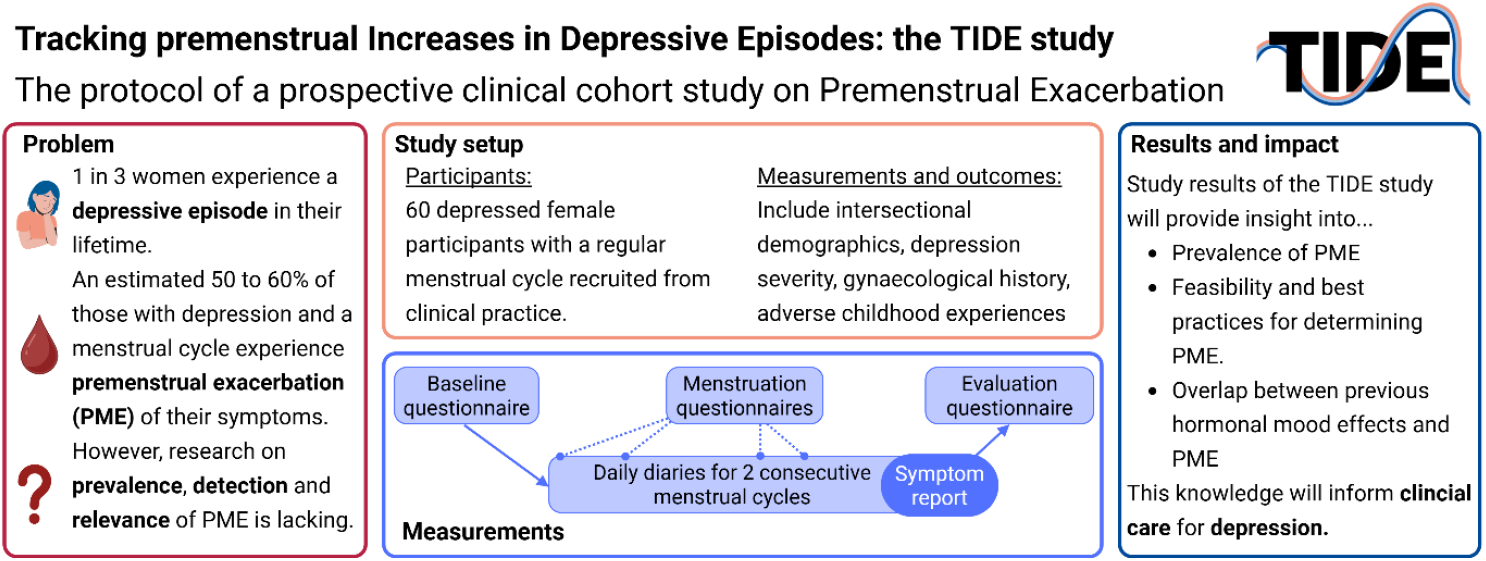

## 1. Introduction

An estimated one in three women in the Netherlands will suffer from depression in their lifetime (Ten Have et al., 2023). There is a growing body of research showing that sex hormone changes could increase the risk of depression or increase the severity of depressive symptoms (Kundakovic & Rocks, 2022), and there is increasing knowledge about mood-related side effects of hormonal contraceptives, premenstrual mood disorders, and perinatal depression. Unfortunately, the role of the menstrual cycle in those diagnosed with a depressive disorder is still understudied.

There are phenomena that describe ways in which the menstrual cycle can affect mood, which all fall under the umbrella of premenstrual mood disorder (PMDs). These include Premenstrual Syndrome (PMS), premenstrual dysphoric disorder (PMDD), and premenstrual exacerbation (PME); an illustration of these phenomena is also visualized in Figure 1. PMS is a phenomenon in which people experience physical and mental complaints in the 7 to 10 days preceding menstruation. Definitions of PMS differ, with some viewing it as a mild everyday inconvenience and others as a clinical phenomenon (Freeman, 2003; Lee, 2002; Walker, 1995). PMDD is a mood disorder in which individuals experience a sudden onset of severe mood symptoms in the premenstrual week, but these symptoms are absent outside of the premenstrual week, with an estimated 3-7% of menstruating individuals suffering from PMDD (Reilly et al., 2024).

**Figure 1.**
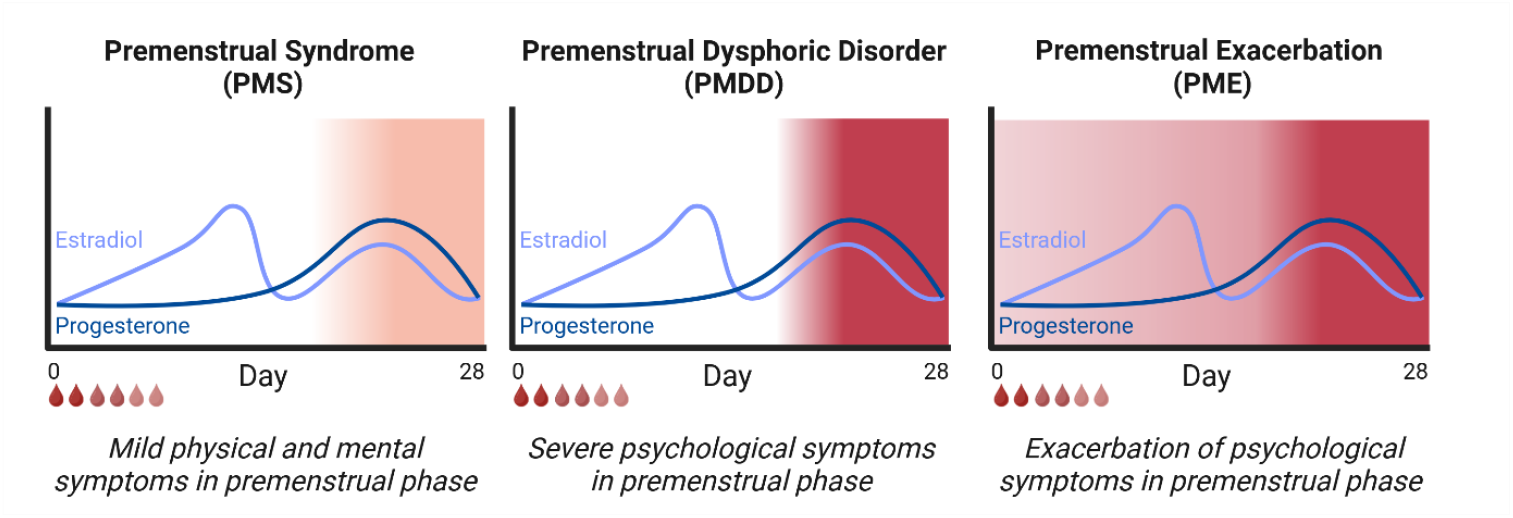
Illustration of symptom severity along the menstrual cycle in premenstrual disorders, including PMS, PMDD, and PME. The color gradients display symptom severity, with mild premenstrual psychological symptoms in PMS, severe premenstrual psychological symptoms in PMDD, and continuous psychological symptoms in PME, which exacerbate in the premenstrual week.

Importantly, one of the diagnostic criteria of PMDD is that the symptoms cannot be caused by existing psychiatric disorders; therefore, someone diagnosed with a depression cannot be diagnosed with PMDD (American Psychiatric Association, 2013). People who experience increases in psychiatric symptom severity in the premenstrual week can, however, suffer from PME, meaning (some of) their symptoms can exacerbate in the 7 to 10 days before the onset of menstruation (Kuehner & Nayman, 2021). PMDD and PME are now commonly seen as similar phenomena that both sit on a continuum of “hormonal sensitivity”, describing how some individuals might be sensitive to the effects of sex hormones on mental health (Pope et al., 2017). This sensitivity could result in adverse mood changes in reaction to the sex hormone changes in the menstrual cycle. The theoretical framework of this hormonal sensitivity to hormonal effects on mood was recently expanded and discussed at length in a paper by Peters et al. (2025), who outlined the DASH-MC framework, a transdiagnostic framework incorporating the menstrual cycle in mental health disorders.

Research into PMDs shows how some people can experience a strong effect of the menstrual cycle on their mental well-being. Studies in people with PMDD show that the premenstrual symptoms, which include anhedonia, low mood, anxious feelings, or feeling overwhelmed, mood lability, irritability and anger, can be severe and have a large impact on sufferers’ mental health (Brown et al., 2024). Symptoms of PMDD show a striking overlap with depressive symptoms, and it is likely that some people with existing mood disorders can experience a similar effect of the menstrual cycle on their mental health, but studies on PME in depression are still scarce.

### Studies on PME in depression

Thus far, there have been a few studies on the prevalence of PME in depressed cohorts. Both Hsiao et al. (2004) and Kornstein et al. (2005) examined PME, using clinical interviews with one retrospective question on experiencing PME. Hsiao et al. (2004) found that out of 50 menstruating patients with a depressive disorder, 80% retrospectively reported PMS, and 52% reported PME of depression. Kornstein et al. (2005) examined the prevalence of PME in the first 1500 participants of the STAR-D cohort, found that 64% of respondents retrospectively reported PME of their symptoms, and that participants who retrospectively reported PME also reported a longer length of the current depressive episode compared to participants who did not report PME (30 vs. 13 months). Subsequently, Haley et al. (2013) used a bigger cohort of the STAR-D to examine PME, finding that those with PME showed a higher prevalence of previous depressive episodes (80% vs. 71%), a higher rate of family history of depression (64% vs. 56%), a higher number of lifetime depressive episodes (5.2 vs. 4.4), and again, a longer duration of the current depressive episode (24 vs. 19 months).

One significant limitation in these retrospective studies is that retrospective symptom reporting in PMDs has been shown to be relatively unreliable. Assuming that studies on PMDD are mostly comparable to PME, there is good reason to suspect that retrospective questionnaire methods are not sufficiently valid to assess the presence of PME. To date, two studies (i.e., Mirghafourvand et al., (2015) and Henz et al. (2018)) compared the prevalence of PMDD as measured using the retrospective premenstrual symptom screening tool (PSST) questionnaire, and compared these rates to prospectively tracked symptoms using the Daily Record of Severity of Problems (DRSP). Results show overestimation in the retrospective PSST compared to the prospective assessment using the DRSP for both severe PMS (42% vs. 16%) and PMDD (35% vs. 4%). The large discrepancies between retrospective and prospective ratings highlight the importance of prospective daily symptom tracking for sufficient symptom insight into PMDs.

Only two studies to date used prospective methods to study PME in clinically depressed participants. Hartlage et al. (2004) asked 900 female participants, of whom 842 were non-depressed, 26 subclinically depressed, and 32 depressed, to track daily symptoms and their mood. They then compared symptom severity between the mid-follicular phase days (i.e., cycle day 5 to 10) and the 7 premenstrual days (i.e., cycle day -7 to -1). Findings show that 58% of the depressed participants reported at least one symptom worsening in the week before menstruation. Delray et al. (2025) examined mood and energy levels in 352 depressed women in a daily dairy study. The authors found that daily mood deteriorated significantly from 14 days before menstruation onset until 3 days after menstruation onset, with 54% of participants reporting a signficantly lower mood in this period. However, Delray et al. did not examine the feasibility or clinical relevance of detection of PME in their cohort.

### The current study

There are no diagnostic guidelines or methods for PME, while PME is associated with a longer recovery time and significantly higher risk of recurrent depressive episodes. Therefore, we have started the TIDE study, in which we strive to improve insights into the role of the menstrual cycle in depression. The TIDE project has the following research questions:

1.Is it feasible and useful to use daily tracking for the diagnosis of premenstrual exacerbation of depression in a clinical cohort?

2.What is the prevalence of PME when assessed using a daily diary screening method (i.e., the golden standard detection method)?

3.Do patients with PME of depressive symptoms also retrospectively report higher rates of other hormone-related mood symptoms, including adverse mood on hormonal contraceptives or peripartum depression?

Altogether, the TIDE study aims to examine the detection, prevalence, and relevance of detecting PME within clinical care for depression.

## 2. Methods

### Recruitment

In the TIDE study, we aim to include 60 female patients who are naturally cycling, diagnosed with unipolar depression and who are currently in treatment in an outpatient mental healthcare setting. Patients are eligible for participation if they are female, between 18 and 45 years old, have a regular menstrual cycle, and are not on hormonal contraceptives. Patients are not eligible for participation if they do not have a good command of the Dutch language, if they do not own a smartphone with internet access, if they are too severely burdened by their psychiatric symptoms to participate in a diary study, if they started menstruating less than 3 years ago, if they are actively trying to get pregnant, if they are pregnant, if they were pregnant less than a year ago, if they are breastfeeding, or stopped breastfeeding less than a year ago. Furthermore, participants with bipolar disorder are also not eligible for the current study, due to unique clinical presentations of PME in bipolar disorder, which can also include manic or psychotic symptoms (Teatero et al., 2014). Due to the observational nature of the study, participants can partake, regardless of any therapy or medication.

Eligible patients are approached either via their healthcare provider or via online routes (i.e., patient foundations, social media). Via a custom QR code and secure contact form, patients can sign up to be contacted by the research team, and the research team will contact them within 5 working days. Patients are then informed about the study by a research team member and receive the information sheet of the study. If they are still interested in study participation, the research team member sets up a second appointment to conduct remote inclusion into the study. During this second appointment, the patient is able to ask questions, discuss the study, and sign informed consent. The first participant in the TIDE study was recruited on August 15^th^ 2025, and the recruitment process is currently ongoing.

### Procedures

Participants will start study participation after signing the informed consent. The start of the study can happen at any point in the menstrual cycle, but study participation ends after the participant has recorded at least two sets of premenstrual and follicular phase weeks; for an additional illustration, see Supplementary Figure 1.

This study has been approved as a non-WMO study (nr. 2025.0018) by the Medical Ethical Committee of the Amsterdam UMC. Due to the observational non-interventional setup of the study, a data monitoring committee is not necessary, as is auditing. We will notify the Medical-Ethical Committee of any significant protocol changes or amendments.

### Measures

Participants provide data in four different types of questionnaires. Below, we explain the timepoint and content of each questionnaire. Please note that all study materials are available in full on the study OSF page (osf.io/4f2y8).

#### 1. Baseline questionnaire

The baseline questionnaire inquires about patient demographics, mental health diagnoses and history, depressive symptom severity, childhood trauma, social support, reproductive history and history of premenstrual symptoms, hormonal contraceptive use, and hormonal medication experiences, and perinatal depression history. This set of measures enables us to assess the diversity of the sample and conduct intersectional analyses, and to account for possible risk factors (comorbidity, childhood trauma) as well as protective factors (social support).

Furthermore, the inclusion of experiences with hormonal medication and pregnancy enables us to examine whether adverse experiences during other hormonal fluctuations are more prevalent in participants with PME. The full content of the baseline questionnaire, including instruments, is shown in Supplementary Table 1.

#### 2. Menstruation symptom questionnaire

Since the TIDE study aims to examine how to best detect PME in clinical practice, we have opted to also examine premenstrual symptoms repeatedly at two time points in the cycle. On day 1 and 10 of the menstrual cycle, participants will be prompted to receive the PSST, which will be modified to ask how participants have been feeling in the past 7 days, reflecting premenstrual symptoms on cycle day -7 to 1 as well as premenstrual symptoms on day 3 to 10, which aligns with PMDD diagnosis criteria (C-PASS). The inclusion of repeated PSST assessments will enable us to examine whether this method could be a suitable and more accessible screening instrument for PMDD or PME in clinical practice.

#### 3. Daily diaries

After inclusion into the study, participants will complete daily diaries on their smartphone. The diary questionnaires are sent every day, and participants will be instructed to set a reminder alarm for the questionnaire at a time around their regular bedtime. The diaries will ask about menstruation (occurrence, physical complaints), depressive symptoms, PMDD symptoms, and whether the participant had unplanned contact with a healthcare provider. The diary items assessing depressive symptoms and PMDD symptoms have been selected from the PHQ-9 and the DRSP to reflect the severity of key symptoms of depression and PMDD, as based on the DSM-5. Due to the length of the DRSP, we have opted to only include the core emotional and physical symptoms of PMDD. Furthermore, due to overlap between the DRSP and PHQ-9, we opted to omit overlapping symptom items to limit the diary length. Individual items are listed in Supplementary Table 1, and the full diary is available on the project OSF page (osf.io/4f2y8).

#### 4. Study evaluation questionnaire

After completion of at least two menstrual cycles, the research team puts together a report for every participant to show the menstrual symptom course, and which symptoms show premenstrual exacerbation. Ten days after sending this study report to the participant, the team sends a subsequent study evaluation questionnaire about how the participant experienced participation in the study (i.e., perceived burden and usefulness of the study diaries), their opinion about the study report (i.e., content and presentation of the report, usefulness), and how they used the report results in their lives and treatment.

### Sample size

We aim to recruit 60 participants into the study. Based on an estimated prevalence of 60% of participants experiencing PME of their depression, this would result in a sample of 36 participants experiencing PME and 24 participants who do not experience PME of their depression. This sample was determined based on the first and second study objectives. Considering the first objective (i.e., feasibility of daily tracking), a sample of 60 participants would give us a first insight into the feasibility and burden of study participation based on participant’ evaluation questionnaires. Considering the second objective of our study (i.e., prevalence of PME), we aim to quantify the extent to which participants have PME based on several methods (see Analysis section), of which most validated and most used method is the C-PASS scoring method. The C-PASS scoring method was also applied in a cohort of 15 unmedicated women with bipolar disorder (Eisenlohr-Moul et al., 2018), which shows that this method also works for small cohorts.

### Data management

All data will be saved and analyzed in the secure environment of the Amsterdam UMC or a similarly secure cloud service provided by the Amsterdam UMC. The only people with access to the raw data are the research team, and participants’ healthcare providers will not have access to the study data. The study report showing the summary of the daily diaries will be sent directly to the participants, and it is up to the participant to decide whether they want to share this report with others, including their healthcare provider.

### Analyses

This study has three main aims, and below we will outline how we will analyze the outcomes for each aim. Please note that the categorization of follicular vs. premenstrual symptom severity is also visualized in Figure 2.

**Figure 2.**
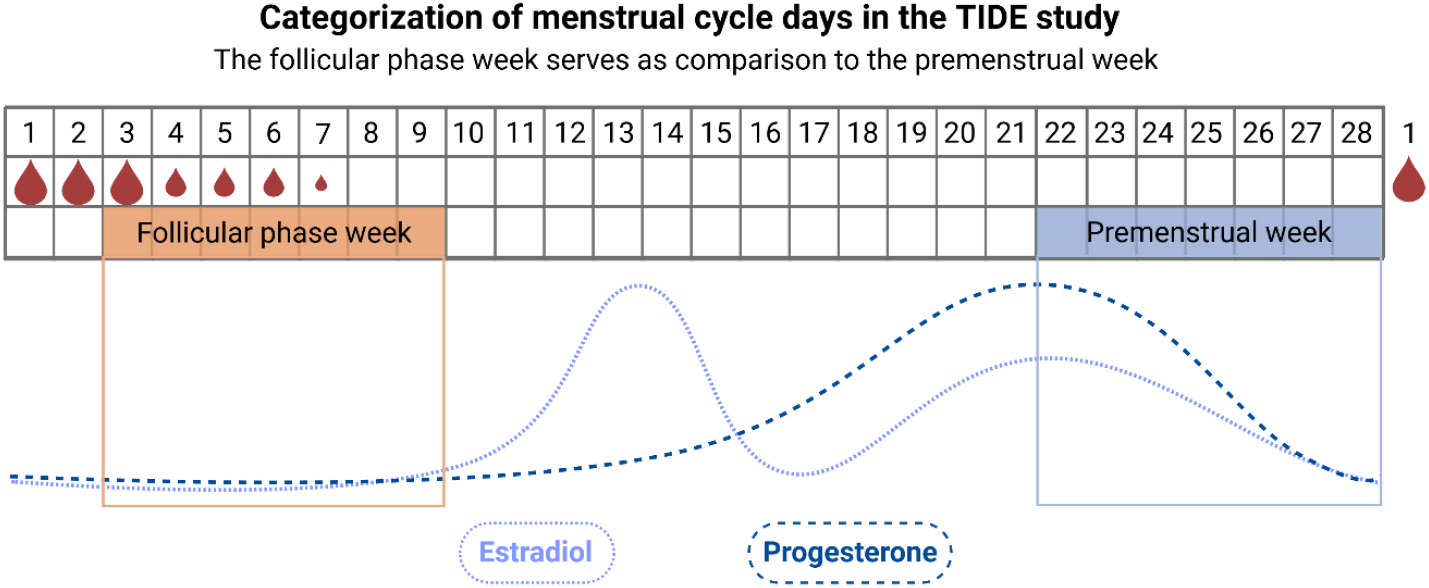
Categorization of follicular- and premenstrual-phase diaries, alongside the hormone fluctuations of the menstrual cycle. As shown in the hormone fluctuations, the follicular phase features low estradiol and progesterone levels, whereas the premenstrual week features decreasing estradiol and progesterone levels.

#### 1. Feasibility and usefulness of daily tracking to measure PME

The first aim of this study is to determine the feasibility and usefulness of daily diary tracking to determine the extent to which participants experienced PME. We will determine the feasibility based on the drop-out rates and the proportion of missing diaries, combined with the reported experiences from participants in the study evaluation questionnaires. We will also determine the usefulness of daily tracking based on the study evaluation questionnaire, in which participants report to what extent they found study participation useful, and how they used the summary of their symptoms in the study report. The study evaluation questionnaire will also contain several open-ended questions and text boxes, which we will analyze for consistent themes.

#### 2. Determine the prevalence of PME in a depressed clinical sample

The second aim of this study is to determine the prevalence of PME of depressive symptoms in a naturally cycling clinical sample in treatment for depression. Since there is no decided clinical or scientific threshold or scoring method thus far to determine whether someone suffers from PME of their depressive symptoms, we will use several scoring methods to assess the presence of PME in depression and report the discrepancies between each method. We outline each scoring method below:

- *Effect size method*: This method was developed by Schnurr (1989) and used by Eckerd et al. (1989), to account for the overall variability in symptoms during the menstrual cycle. In the effect size method, the “effect size” is an indicator of how much symptom severity varies between the premenstrual and follicular phases, while considering overall variability. The effect size is calculated by subtracting the average symptom rating in the luteal phase from the symptom score in the follicular phase. We adapted these time categories to align with the C-PASS method, comparing average symptom severity in the seven premenstrual days (days -7 to -1) with seven follicular phase days (days 3 to 10). The difference between the average luteal and follicular phase is then divided by the standard deviation (SD) of the ratings in the full menstrual cycle. The resulting score represents the degree of PME, with a higher score representing more severe PME. A score of 1.0 or higher has been considered a marker of the presence of PME (Hartlage et al., 2012). To categorize the presence of PME in the cohort, we will report how many participants show PME in a depression core symptom (i.e., depressed mood or anhedonia), and how many participants show PME in 3 or more symptoms.
- *C-PASS*: Following the C-PASS criteria, developed by Eisenlohr and colleagues (2017; see Box 1), we will examine how many participants meet the diagnostic criteria for PMDD or menstrual-related mood disorder (MRMD). This will show how many participants show a relative absence of their symptoms after the onset of menstruation, indicating that there is possible clearance of symptoms in the follicular phase. It is important to note that that the daily diaries of the TIDE study contain a selection of the DRSP items but not all items, meaning we cannot completely follow the C-PASS scoring, and we will omit the criterion inquiring about the impact on daily life.
- *PACTS:* All aforementioned methods rely on phase-based categorization of diary outcomes, thereby possibly oversimplifying the menstrual-cycle related changes in symptoms. In a recent study, Nagpal et al. (2025) described the Phase-Aligned Cycle Time Scaling (PACTS) method, with corresponding software package, providing researchers with continuous time variables to study the menstrual cycle, which are centered around the onset of menstruation or ovulation. Within the TIDE study, we will use the PACTS method to examine continuous associations between depressive symptom severity and time from menses onset.

As a secondary outcome, we will also use the retrospectively reported PSST scores from the baseline questionnaire and the PSST scores from day 1 and 10 of each cycle to compare how retrospective scoring of menstrual symptoms compares to prospective scoring of symptoms.

#### 3. Overlap with symptoms during hormonal transitions

The third aim of this study is to examine whether participants who experience PME also retrospectively report a higher rate of depressive symptoms around previous hormonal changes. To meet this aim, we will compare the prevalence of perinatal depression (reported using the Lifetime Edinburgh Postnatal Depression Scale; Meltzer-Brody et al., 2013) and the prevalence of mood-related side effects of hormonal contraceptives between participants who have no PME or in whom there is mild or severe PME. We will compare these groups using binomial linear regression analyses, taking into account other risk factors for depression (e.g., socioeconomic status, religious, ethnic or LGBTQ minority status, childhood trauma, and social support).

### Adherence

One of the risks of this study, especially in participants with PME, is specific non-adherence or missing data in the study diaries in the weeks or periods when participants are feeling worse, possibly resulting in undersampling during the premenstrual phase. Therefore, we aim to optimize adherence in two ways, namely through personal rapport building and participant involvement in the outcomes. Firstly, each TIDE study participant will have one specific research team member with whom they will have primary contact and who will be available for questions or concerns during study participation. The research team will check adherence to the diaries, and the research team member will check in with participants if they miss several consecutive diaries. Second, participants will receive a report of their own measurements after they complete the study diary period, which can offer them insights about the role of the menstrual cycle in their own mental health. The research team will emphasize the importance of completing the diaries, even on days when participants feel poorly, to ensure that the report is as relevant, useful and realistic as possible.

Nonetheless, it is still possible that participants will drop out or will not complete follow-up; therefore, participants who discontinue participation will still be asked to fill out the study evaluation questionnaire to get better insights into their experiences participating in the study. We will also examine whether we have selection bias in loss to follow-up by comparing demographic characteristics between participants who are or are not lost to follow-up.

### Open science and dissemination

Due to the sensitive nature of the data, we will not share the raw study data publicly; instead, collaborators or interested parties who are interested in using the study data can apply to use the study data by writing a paper proposal. After approval of the paper proposal by the study leader, logistics for data sharing will be arranged, either through a data sharing agreement or through the invitation of the collaborator to a secure cloud server through which they can access the data. We will upload time-stamped versions of the study protocol and study questionnaires, as well as any relevant analysis scripts, to the study OSF page (osf.io/4f2y8) after the publication of the first study results. The analyses for the current study aims were not preregistered, but we will preregister analyses which fall outside of the scope of the original study aims.

All study results will be disseminated via the study website (www.TIDEstudy.com), as well as via academic publications. Dissemination to non-academic parties (i.e., healthcare providers, stakeholders, general public) will happen through presentations at conferences, collaboration with relevant patient organizations (i.e., PMDD Netherlands), and by giving presentations to local clinical teams.

## 3. Results

Daily diary tracking is a powerful tool to improve self-insight into symptoms, and research increasingly shows how self-reported diary assessments are deemed important and useful for clinical care. Therefore, we opt to provide participants who completed sufficient daily diaries (i.e., at least 70% of the sent diaries in each menstrual cycle) with a report displaying their own diary responses. This report was set up with input from several stakeholders who live(d) with depression, PMDD, and PME, who provided input in a shared brainstorm about what the diaries should report. After this brainstorm, we sent around a preliminary version of the diary report, and the stakeholders provided feedback on a preliminary version of the report, which resulted in the final version of the report. The final version of the diary report contains a short explainer about what the menstrual cycle is and then shows the average symptom burden and specific symptom burden per cycle day, as well as which symptoms significantly increased in the week before the onset of menstruation.

### Symptom report for participants

The figures summarizing the participants’ responses in the report are created using RStudio, using standardized code written by authors IdV and MM (full code can be found on osf.io/4f2y8). Figure 3 below shows an example of a graph included in the participant report.

**Figure 3.**
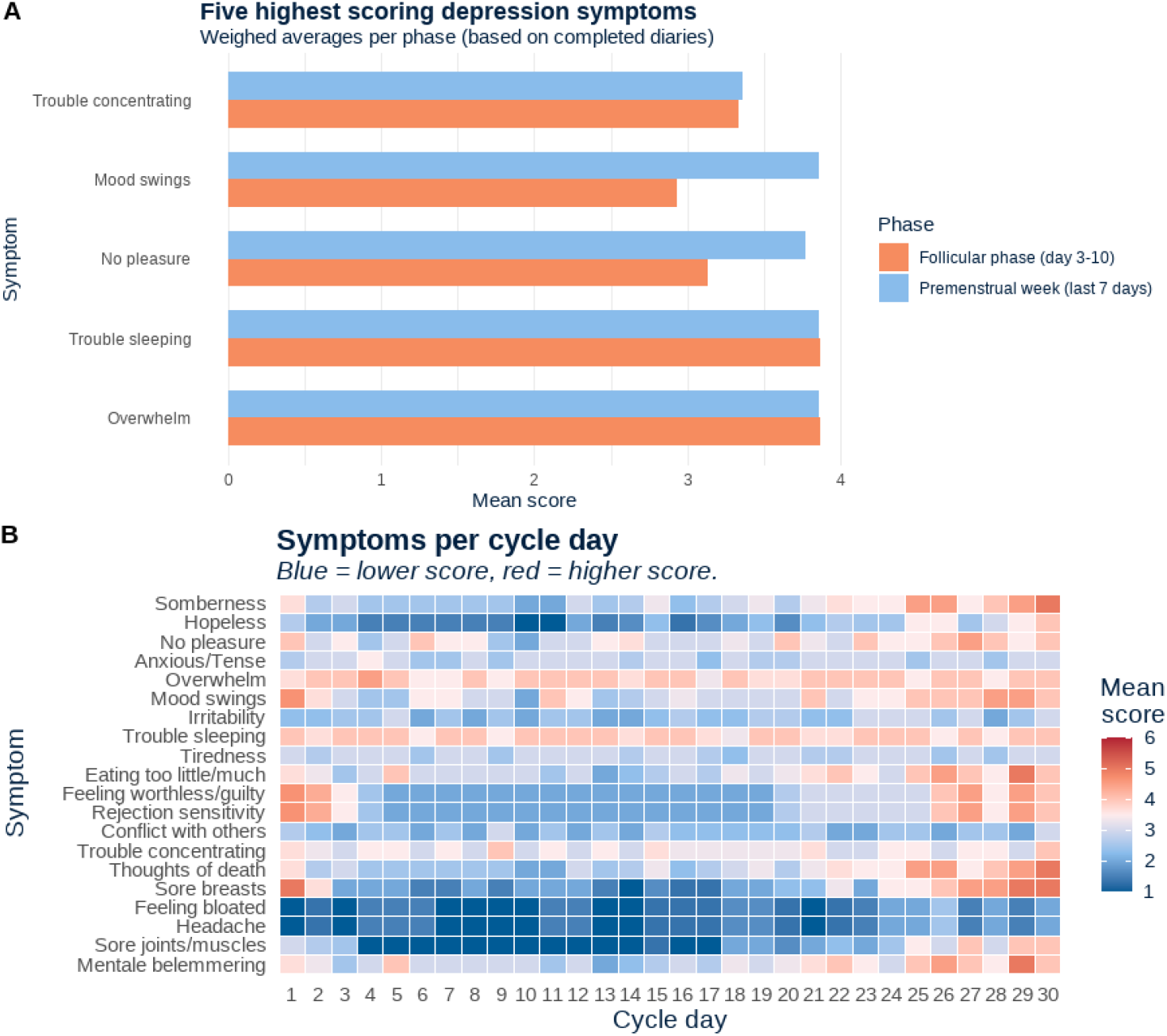
**Panel A:** Bar plot displaying the top 5 most severe symptoms recorded across the measurement period, stratified by menstrual cycle phase. **Panel B:** Heat map displaying depression symptoms and somatic premenstrual symptoms across the menstrual cycle for a hypothetical participant. The hypothetical participant shows premenstrual exacerbation in somberness, hopelessness, and sore breasts.

The full symptom report is sent to participants directly, and participants are told that they are free to show the report to whomever they want. In the study evaluation questionnaire, participants are asked whether they showed the report to loved ones or healthcare providers, whether they found the report useful, what aspects they found most useful and whether they have feedback for the research team to improve the study or symptom report. Although the diary report is not part of the primary aims of this study, the participant feedback can provide insight into whether a the symptom report is beneficial for patients with a depression and possible PME.

## 4. Discussion

The aims of the TIDE study are to establish the feasibility and relevance of measuring PME of depressive symptoms in a clinical cohort, to estimate the prevalence of PME in a depressed clinical cohort, and to examine associations between PME and other hormone-related depressive symptoms. Previous studies have retrospectively or prospectively tracked PME in depression, but these previous studies are scarce, most used retrospective methods, and no study asked participants themselves to report the feasibility and usefulness of the day-to-day symptom tracking. Furthermore, authors of the one previous study collecting prospective data did not yet compare the different scoring techniques of the daily data. Additionally, our study will add knowledge on the possible overlap between sex hormone-associated depressive symptoms, by also examining retrospectively reported perinatal depression and hormonal contraceptive-associated side effects.

### Benefits of increased insights into PME

Although the requirement of daily symptom tracking in assessing PME could be burdensome on patients, daily tracking also offers important opportunities and insights for those with PME and their healthcare providers. First, daily symptom tracking could offer improved insights into symptom trajectories over time, enabling patients to recognize the cyclical nature of the symptom surges. Qualitative studies in those with PMDD repeatedly find that daily symptom tracking is an important step for recognition and awareness of the symptoms (Brown et al., 2024), helping those with premenstrual mood disorders feel more in control of their mental health. In care for depressed patients, the recognition of symptom surges could also help patients’ social contacts and healthcare providers, since it could help them anticipate more difficult (i.e., premenstrual) weeks in which symptoms might increase. Secondly, recognizing a PME-like pattern in symptom trajectories could also offer opportunities to discuss additional treatment options. These range from increased support in the premenstrual week (i.e., extra contact with HCPs or social support from loved ones), to risk assessment (i.e., insight in self-harm, thoughts of death or need for urgent care in the premenstrual week), to trying additional treatment strategies to reduce cyclical worsening (i.e., adapting medication dosage in the premenstrual week or starting hormonal contraceptives, based on PMDD treatment options; Carlini et al., 2022), although we must note that there are no existing treatment guidelines for PME in depression. Lastly, experiencing PME could also be a marker of hormonal sensitivity, meaning someone could be especially sensitive to mood symptoms during sex hormone changes (Pope et al., 2017). This “hormonal sensitivity” is a hypothesis currently being studied by multiple research groups, and the first studies have found that those with severe PMS or PMDD are also more likely to experience adverse mood in the peripartum period (Pereira et al., 2022). Therefore, PME could also be a marker for risk of future depressive symptoms during sex hormone changes.

### Conclusion

To our knowledge, this is the first study to combine daily clinical symptom tracking and participant feedback in examining PME of depression in a clinical cohort. We expect this study to provide unique insights into experiences with PME in a cohort of depressed patients, which can lead to several new clinical and fundamental studies into the role of the menstrual cycle in depressive symptoms. This could include novel treatment studies working towards additional treatment options in patients with PME: once the prevalence and detection method for PME is determined, we could examine whether luteal- or premenstrual-phase increases in antidepressant dosages (Miller et al., 2008) or addition of oral contraceptives to treatment (W. Peters et al., 2017) could be acceptable and effective strategies for reducing PME. This could offer new, personalized treatment strategies for depression, working towards better outcomes, and personalized medicine in mood disorders.

## Data Availability

The manuscript does not describe original data. The study protocol, materials, data, and analytic code are available via osf.io/4f2y8.

## Acknowledgements

The authors wish to thank all the volunteers and experts with lived experience from Stichting PMDD Nederland, who contributed their insights, ideas, and feedback to the TIDE study. Furthermore, the authors wish to thank the clinicians who helped set up the study or contributed to recruitment for the TIDE study.

## Author contributions

Chamee Giezenaar: *Investigation; Resources; Writing – Review & Editing; Project administration*. Isis de Vries: *Software; Investigation; Resources; Data Curation; Writing – Review & Editing; Visualization*. Margot Morssinkhof: *Conceptualization; Methodology; Data Curation; Writing – Original Draft; Visualization; Supervision; Funding Acquisition*.

