## Supplementary materials for "Tracking premenstrual exacerbation (PME) of depression in a prospective clinical cohort: the TIDE study protocol"

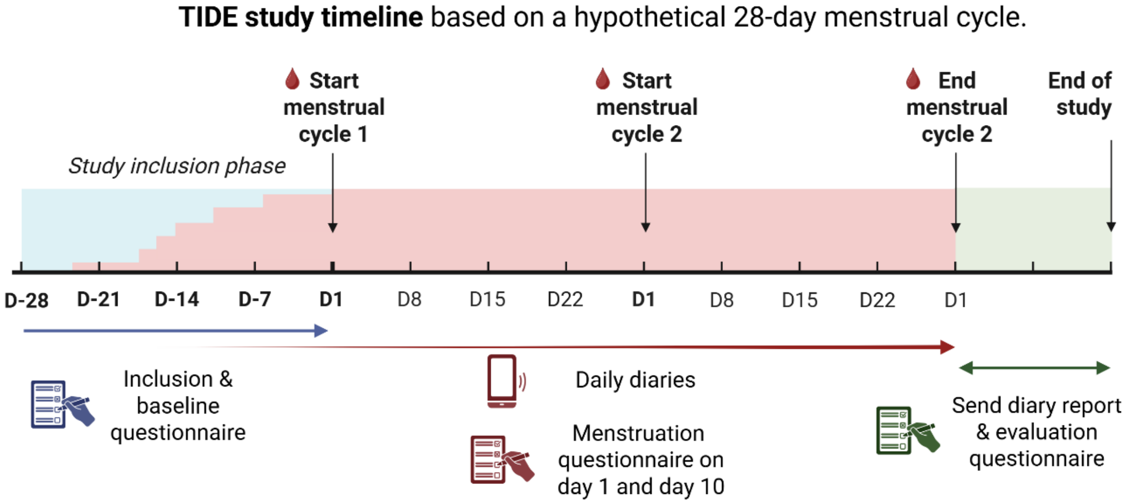


**Supplementary Figure 1.** Timeline of study participation in the TIDE study. Participants complete an inclusion questionnaire, daily diaries for two consecutive menstrual cycles, menstruation questionnaires of day 1 and 10 of the menstrual cycle for two consecutive cycles, and a study evaluation questionnaire.

| Supplementary Table 1. Study questionnaires, outcomes and instruments. | | |
|---|---|---|
| **Measure and time** | **Outcome** | **Instrument** |
| **Baseline questionnaire – asked at start of daily diary tracking** | Demographics | Age, gender, sexual orientation, relationship status, cohabitation status, migration status, ethnicity, education level, work status, financial status |
| | Mental health history | Previous diagnoses, current diagnoses, depressive episode (first vs. recurrent), age at first depressive episode, medication use (form, dose, duration) |
| | Depressive symptom severity | CES-D (Radloff, 1977) |
| | Childhood trauma | SF-CTQ (Bernstein et al., 2003; Thombs et al., 2009) |
| | Social support | ENRICHD Social support instrument (Mitchell et al., 2003) |
| | Reproductive history | Age at first menses, menses regularity, cycle duration and use of cycle tracking, previous pregnancies and live births |
| | Experiences with hormonal contraceptives | Self-constructed, including:<br>Use of hormonal contraceptives, age at start, duration of use, reason for start, reason for cessation, side effects, rating of contraceptive (1-10) |
| | Retrospectively reported PMS or PMDD | Premenstrual Symptom Screening Tool (Steiner et al., 2003) |
| | Lifetime history of perinatal depression | Lifetime Edinburgh Postnatal Depression Scale (Meltzer-Brody et al., 2013) |
| **Menstruation questionnaire – Cycle day 1 and 10** | Premenstrual symptoms in past 7 days | Premenstrual Symptom Screening Tool [edited to reflect past 7 days] (Steiner et al., 2003) |
| **Daily diaries – Daily for two consecutive cycles** | Day rating | Grade on a scale of 1 to 10 |
| | Menses | Occurrence of menstruation or spotting<br>Physical menstruation symptoms and symptom burden |
| | Key PMDD symptoms | DRSP items 1, 2, 3, 4, 5, 6, 7, 8, 16. |

| | | |
|---|---|---|
| | Physical PMDD symptoms | DRSP items 18, 19, 20, 21. |
| | Depressive symptoms | PHQ-9 items 1, 3, 4, 5, 7, 9 |
| | Urgent mental health care | Unplanned contact with healthcare provider, use of additional medication (i.e., anxiolytics, dose increase) |
| **Study evaluation questionnaire** | Daily questionnaire burden | Diary bother, diary usefulness, worth the time and effort, feasibility, remarks |
| | Usefulness of summary report | Clarity, usefulness, which outcomes were good to see, which outcomes did you miss |
| | New insights from summary report | New insights into symptom trajectories, results relevant for everyday life, life changes based on report, remarks |
| | Role of healthcare providers | Shared report with healthcare provider, healthcare provider reactions, changes in treatment plan |
